# Estimating the false positive rate of highly automated SARS-CoV-2 nucleic acid amplification testing

**DOI:** 10.1101/2021.04.25.21254890

**Authors:** Christopher M. Chandler, Lori Bourassa, Patrick C. Mathias, Alexander L. Greninger

## Abstract

Molecular testing for infectious diseases is generally both very sensitive and specific. Well-designed PCR primers rarely cross-react with other analytes, and specificities seen during test validation are often 100%. However, analytical specificities measured during validation may not reflect real-world performance across the entire testing process. Here, we use the unique environment of SARS-CoV-2 screening among otherwise well individuals to examine the false positivity rate of high throughput so-called “sample-to-answer” nucleic acid amplification testing (NAAT) on three commercial assays: the Hologic Panther Fusion®, Hologic Aptima® transcription mediated amplification (TMA), and Roche cobas® 6800. We used repetitive sampling of the same person as the gold standard to determine test specificity rather than retesting of the same sample. We examined 451 people repetitively sampled over 7 months via nasal swab, comprising 7,242 results. During the study period there were twelve positive tests (0.17%) from 9 people. Eight positive tests (0.11%, five individuals) were considered bona fide true positives based on repeat positives or outside testing and epidemiological data. One positive test had no follow-up testing or metadata and could not be adjudicated. Three positive tests (three individuals) did not repeat as positive on a subsequent collection, nor did the original positive specimen test positive on an orthogonal platform. We consider these three tests false positives and estimate the overall false positive rate of high-throughput automated, sample-to-answer NAAT testing to be approximately 0.041% (3/7242). These data help laboratorians, epidemiologists, and regulators understand specificity and positive predictive value associated with high-throughput NAAT testing.

## Introduction

During assay validation, clinical tests are specifically interrogated for their sensitivity and specificity. (1) Sensitivity is defined by the ability of the test to return a positive result in presence of an analyte, while specificity indicates the test’s ability to return a negative result in the absence of that analyte. While both are important, test specificity can be an especially critical parameter for screening tests, where the vast majority of persons are negative. This fact has been well-appreciated in infectious disease serological testing for viruses such as human immunodeficiency virus (HIV) or hepatitis C virus (HCV), but has not been widely interrogated for infectious disease nucleic acid amplification testing (NAAT) in great detail, as the cost of molecular testing has restricted its use in screening. In addition, there is little commercial incentive to advertise a laboratory’s false positivity rate, even though both laboratorians and physicians understand that false positives can occur at any point in the testing process.

The performance of molecular infectious disease testing has generally been evaluated based on analytical sensitivity, since analytical specificities of most NAAT, including polymerase chain reaction (PCR) tests, during validation are nearly always 100%. With the widespread availability of genomic data today, it is generally facile to specifically design PCR primer and probe sets to avoid cross-reaction with other analytes. While several SARS-CoV-2 primer sets were initially designed to specifically cross-react with SARS-CoV or potentially other *Sarbecovirus* members, no primer sets were specifically designed to cross-react with other known circulating human viruses. (2)

Given the long-term shedding associated with SARS-CoV-2 infection (over half of previously hospitalized patients in a multicenter study in the United States had a positive PCR test at 3 weeks or longer following their first positive test) and the superlative analytical sensitivity of NAAT, adjudicating the trueness of low viral load positives can be difficult. (3) Retesting a positive specimen on an orthogonal testing platform may help determine true analytical positivity if the second test is positive, but a negative test may not be specifically informative if the viral load is at the analytical limit of detection (LoD). An alternative method to determine false positivity would be to recollect individuals, though this still has problems with stochasticity of samples, especially at the LoD.

The SARS-CoV-2 pandemic creates an intriguing use case to examine the overall false positivity rate associated with large-scale NAAT, as millions of tests were performed daily across the United States in 2020-21. Many employers such as technology companies or sports teams specifically tested their employees every day or every other day to prevent widespread transmission within their organization. Our clinical laboratory performed almost half of testing within Washington State and offered repeat longitudinal testing to several employer groups that had exceptionally low positivity rates. Here, we use this data to specifically interrogate the false positivity rate associated with high-throughput, so called “sample-to-answer” assays employed in our laboratory: Hologic Panther Fusion®, the Hologic Aptima® transcription mediated amplification (TMA), and Roche cobas® 6800.

## Materials and Methods

### Description of cohort and testing

This study was approved by the University of Washington Institutional Review Board. Three different, high-throughput sample-to-answer platforms for SARS-CoV-2 testing were used during the study period: the Hologic Panther Fusion®, Hologic Aptima® TMA, and the Roche cobas® 6800 assays. The Panther Fusion® assay targets two conserved regions of the SARS-CoV-2 open reading frame (ORF) 1ab gene; amplification of either or both regions produces a presumptive positive test result and amplification of neither target results a negative test result. The Roche cobas® 6800 assay targets two regions of the SARS-CoV-2 genome, the envelope (E) gene and ORF1ab gene. Amplification of both targets results in a presumptive positive test result, while amplification of one of two targets results in an inconclusive result, and amplification of neither target results a negative test result. The Aptima® TMA assay targets two conserved regions of the SARS-CoV-2 ORF1ab gene; amplification of either or both regions produces a presumptive positive test result and amplification of neither target results a negative test result. An in-house laboratory developed test (LDT) for SARS-CoV-2 was also in use during this time but the results from the LDT are not included in the present analysis as it is a not a commercial, high-throughput platform. However, testing on the LDT is incorporated into the statistics regarding testing frequency for transparency and completeness. Cycle time (Ct) parameters for the commercial assays are considered proprietary and not publicly available. However, Ct results are reported on the instrument for positive or inconclusive tests on both the Panther Fusion® and Roche cobas® 6800 assays (Table 1).

**Table 1.**
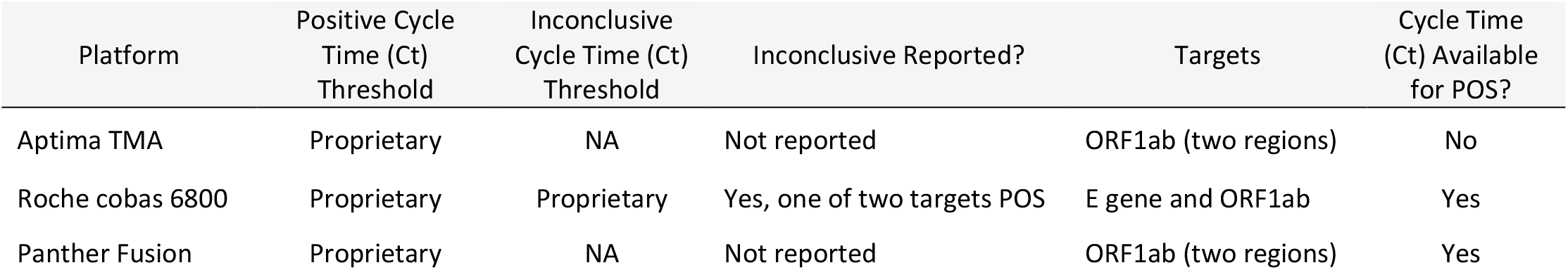
Ct values and targets for each platform

Testing was performed according to the manufacturer’s instructions for use (IFU) as detailed in each respective assay’s emergency use authorization (EUA) through the United States Food and Drug Administration (FDA) and as validated for use in the University of Washington Clinical Virology Lab. The inclusion criteria for samples in this study was any location/employer which had a requirement to confirm any positive result on an orthogonal platform as well as re-collect the patient/employee and re-test within 24 hours to confirm the initial positive result. The specimens were nasal swabs collected at each respective location’s/employer’s facility according to established protocols. Transport to the University of Washington Clinical Virology lab was done in universal transport media (UTM) or viral transport media (VTM). Specimens were processed and run within 72 hours of collection if refrigerated or frozen within 72 hours of collection.

Common preanalytical steps shared by all specimens regardless of assay included aliquoting of the specimen in a biosafety cabinet and transfer of the aliquot to a reaction vessel appropriate for use on the testing platforms. Positive results were deemed “false positives” if the subsequent sample from the same individual was negative, regardless of platform, and, similarly, “true positives” were assigned if the subsequent sample was also positive with a flat or increasing viral load (flat or decreasing Ct). Additionally, positive samples preceded by an inconclusive result were also considered “true positives.”

### Data analysis

Data for the populations of interest was extracted from the Department of Laboratory Medicine and Pathology’s data warehouse that contains patient demographics and laboratory order and result data that are extracted from our laboratory information system (Sunquest Laboratory version 8.1, Tuscon, AZ). Individuals were eligible if they met the above inclusion criteria and had at least two molecular tests for SARS-CoV-2 performed on a high-throughput platform during the study period (May-November 2020). Data analysis was performed using RStudio (Version 1.2.1335) and Microsoft Excel.

## Results

A total of 451 people repetitively tested over a seven-month period from May to November 2020 were included in the analysis. The median age of those tested was 27 years old (interquartile range, IQR, 23-33 years). A total of 7,242 SARS-CoV-2 tests were performed on the high-throughput assays during the study period; all specimens were nasal swabs. The median number of tests per individual was 10 (IQR 6-16) and the median number of days between consecutive tests was 2.1 days (IQR 2-5 days). There were 12 positive (detected) results during the study period (0.17%) from 9 people (Figure 1 and Table 2). There was one inconclusive result (0.013%) from one person and was followed by a positive result. The number of total and positive results by platform were as follows: Panther Fusion®, 1932 (26.7%, six positive); Aptima® TMA, 1526 (21.0%, four positive), and Roche cobas® 6800, 3784 (52.3%, two positive).

**Figure 1.**
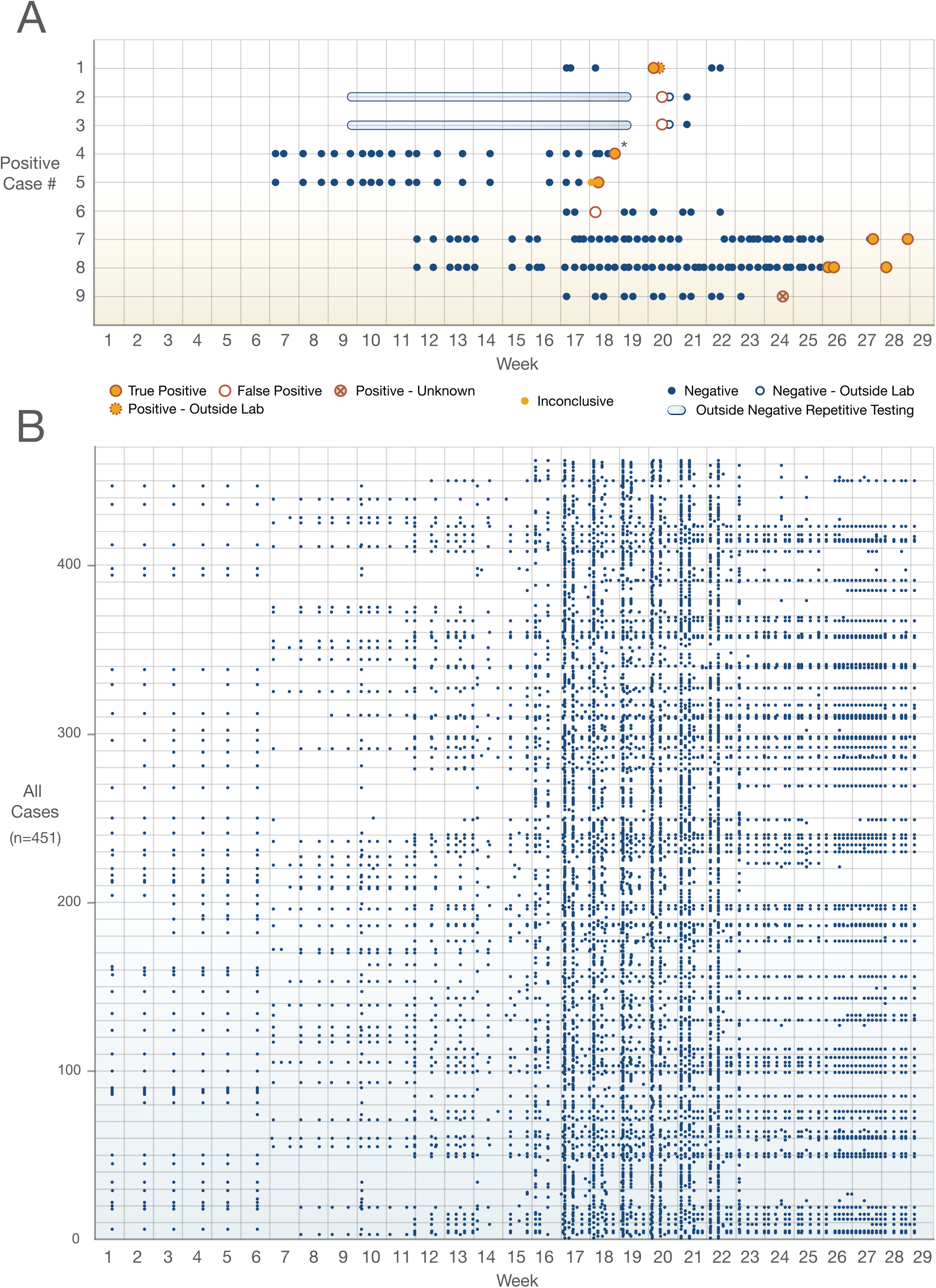
Testing Frequency and Positive Cases The frequency of testing in our cohort during the study period was high with a median of 2.1 days between tests and median of 10 tests per individual. Individuals with a positive (detected) result are broken out in panel A and all results are shown in panel B. Two true positive cases were judged according to outside laboratory data or clinical symptoms; the positive result in Case 1 was followed by a positive repeat test at an outside laboratory a day later and the positive result in Case 4 (*) was interpreted as clinical true positive as the patient developed symptoms within one day of testing and had close contact with a person known to have COVID-19.

**Table 2.**
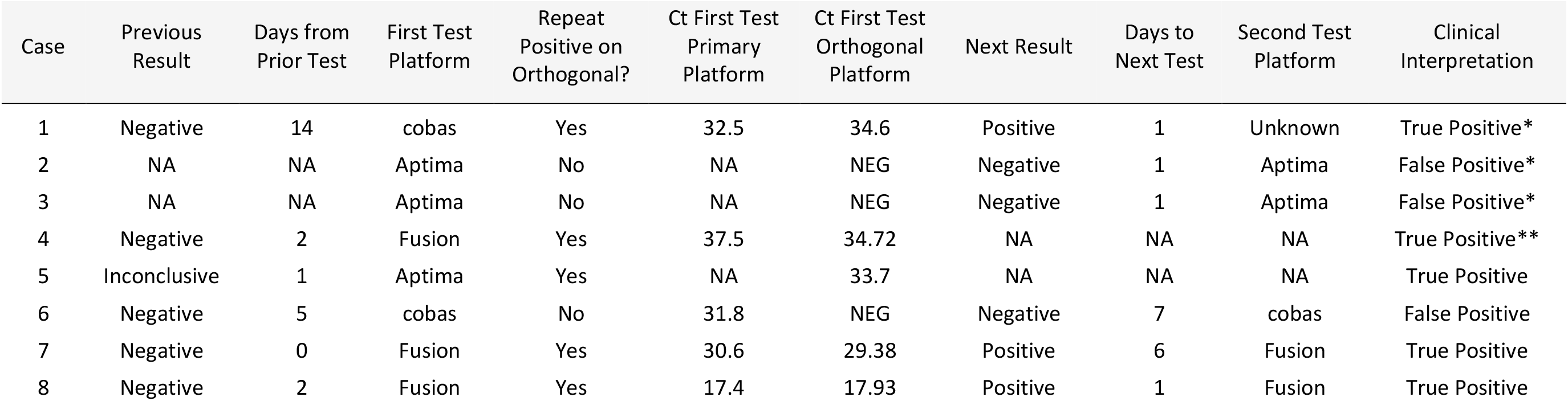
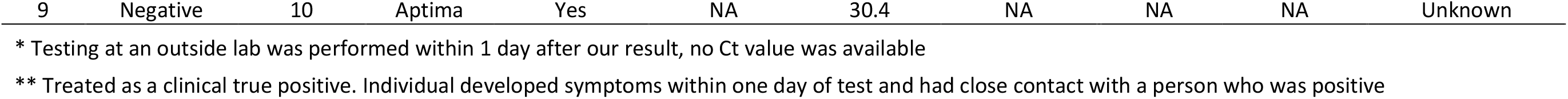
Positive Results and Interpretation

Of the positive results, three were interpreted as false positives as a subsequent collection was negative (Cases 2, 3, and 6). All of specimens which gave false positive results failed to repeat as positive when testing the original parent sample or the aliquot on an orthogonal platform. Only one false positive test had an accompanying Ct value (31.8). The mean time to subsequent collection was 3 days (range 1 to 7 days). Two of the false positive cases (Cases 2 and 3) did not have a preceding result in our system, but did have repetitive testing at an outside lab that was negative per report. We estimate the overall false positive rate of high-throughput automated molecular testing to be approximately 0.041%.

A total of five cases were interpreted true positives. Two positive cases had static or decreasing Ct value on consecutive tests performed in our laboratory (Cases 7 and 8, Figure 2). One case (Case 1) had a positive test result that was followed by a repeat positive test performed at an outside laboratory a day later (no Ct value available). The fourth case (Case 4) was interpreted as clinical true positive as the patient developed symptoms within one day of testing and had close contact with a person known to have COVID-19; no subsequent testing was performed on this individual. Lastly, Case 5 had a positive result following an inconclusive result, which is consistent with detecting an early infection, and we count as a true positive. All of the true positive results were positive using the same specimen on an orthogonal platform. The average Ct value of the five true positives for which this data was available on the primary testing platform was 29.5 and the mean time to subsequent collection was 2.97 days (range 1 to 6 days). Lastly, there was one positive result which cannot be adjudicated as a true or false positive using our criteria as there was no subsequent collection. However, this sample repeated as positive on an orthogonal platform using the same specimen suggesting it may have been a true positive as well.

**Figure 2.**
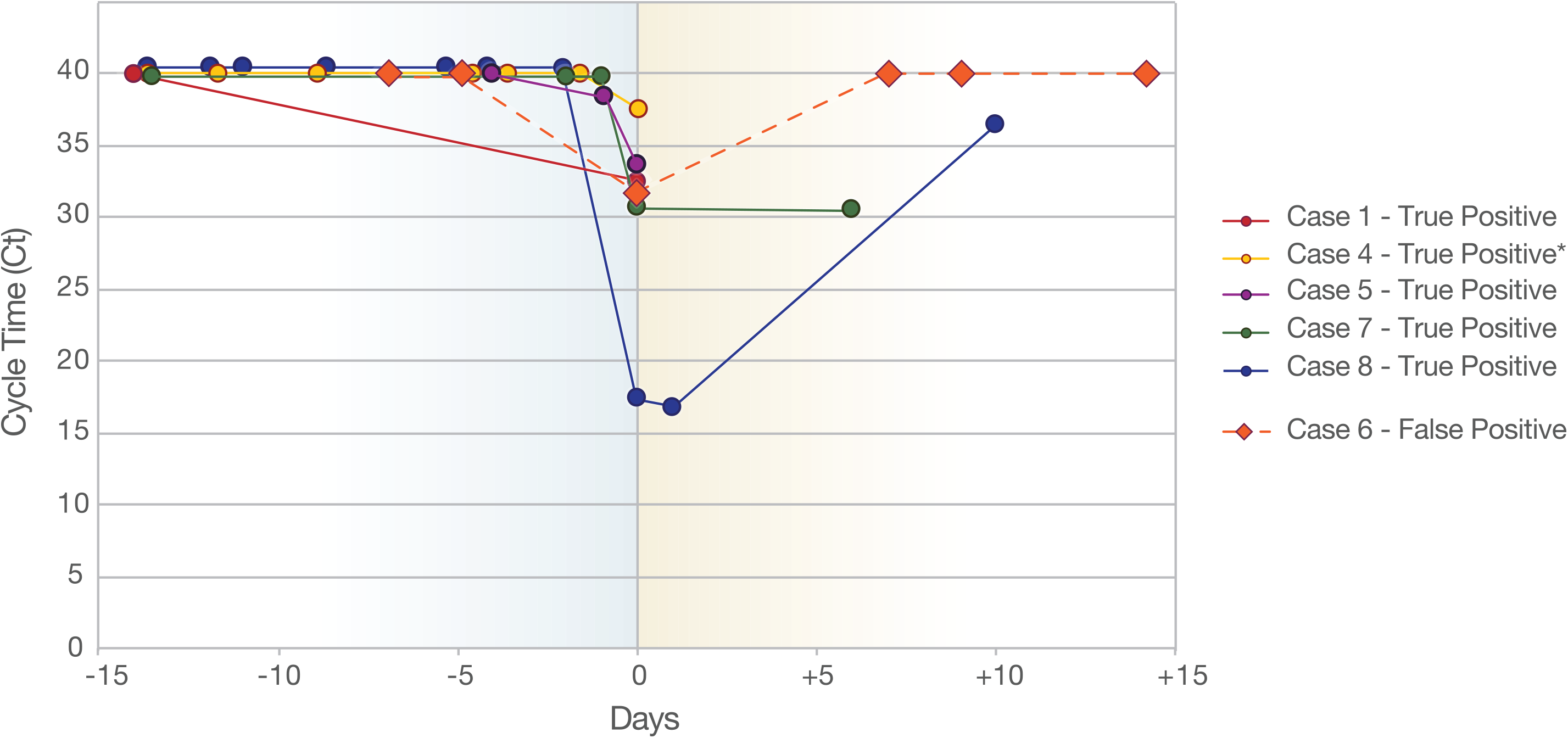
True Positive and False Positives with Ct values over Time Positive SARS-CoV-2 results with available cycle time (Ct) values and follow-up are shown longitudinally in relation to their immediate prior and subsequent results (represented in days since the first positive test, time zero). True positives are characterized by a decrease in Ct value that repeats on subsequent testing whereas false positives show a brief dip in Ct followed by a negative test. A single case (Case 5) had an inconclusive result prior to the positive test (amplification of a single target one day prior) and is considered a true positive of an early infection. All positive results with Ct values on the primary diagnostic platform are included. A Ct value of 40 was assigned to negative results for illustration purpose.

## Discussion

We present an estimate of the false positive rate of high-throughput, so called “sample-to-answer” NAAT for SARS-CoV-2 derived from a healthy, relatively young repetitively sampled population. The overall false positive rate of 0.041% is quite low and concordant with the reported specificity of currently available PCR primers. However, when considering the magnitude of testing for SARS-CoV-2 currently performed on a daily basis in the United States, the number of false positives becomes of greater clinical concern (820 individuals daily if using the figure of two million daily tests in the United States).

It is unclear at which stage of the testing process the false positives we identified originated. The preanalytical stage is most likely as the vast majority of laboratory errors occur during this part of the testing process. (4) Between accessioning and aliquoting into instrument-compatible tubes, the manual manipulation required for testing on automated platforms is still considerable. The analytical stage is also a possibility. As detailed in the study by Lin et al. (2020) false positives can occur during testing due to cross-contamination between neighboring samples when one is a strong positive, resulting in a weak false positive in an adjacent sample. (5) Side-by-side sample cross-contamination was determined to be the cause for 39 of 40 (97.5%) of the false positive results they identified, and led to specific changes in their test procedures including the use of more blanks and reduction in the wash volumes during extraction steps to reduce the risk of splashing. (5) However, their testing was limited to an open, conventional extraction and amplification workflow similar to a laboratory developed test and not the closed, sample-to-answer testing that comprise the results detailed here. Their overall false positive rate of 0.1% is similar to the estimate of 0.041% we obtained.

Additionally, there is the cautionary tale of contamination of test reagents, particularly the non-template controls, by synthetic SARS-CoV-2 oligonucleotides which delayed implementation of RT-PCR testing for the virus in the European Union in January and February of 2020. (6) Similar contamination plagued the rollout of the US Center for Disease Control and Prevention’s (CDC) own SARS-CoV-2 RT-PCR test kits in February and March of 2020 and other instances of contamination of commercial reagents have been published. (7, 8) These examples reinforce the need for stringent laboratory quality control procedures, including routine examination of new lots of test reagent, and vigilance for patterns of positivity which fall outside of expectations (clusters of positive results, side-by-side positives, or any instance of suspected human error during the manual steps of the process).

Another factor are the unique performance characteristics of the assays. Although generally comparable in clinical use, sensitivity and specificity of the tests is not uniform, and they target distinct molecular regions of SARS-CoV-2. (9, 10) Also, the technologies used by each assay are also not the same; the Aptima® TMA assay is an isothermal technology that creates a chemiluminescent signal read by a luminometer, reported in relative light units (RLU), and relies on differences in signal kinetics to generate results whereas the other platforms produce a fluorescent signal and amplify target transcripts using traditional thermal cycling after reverse transcription (RT-PCR). While the Aptima® TMA assay has been reported to be more sensitive than comparable RT-PCR assays, our data suggests this may come at the cost of false positive results. (11) Two out of three of our false positive cases occurred using the Aptima® TMA assay (Case 2 and 3). These positive tests had RLU values of 614 and 615 respectively, which is between the average RLU of 300 for negative samples and 1153 for 0.5xLoD samples in the manufacturer’s IFU, likely indicating that these results were close to the assays LoD.

Lastly, all laboratory test results should be interpreted within the context of their pretest probability of disease. In our cohort of generally well, young individuals tested in the period prior to the recent winter surge of SARS-CoV-2 infections in the United States, the pretest probability for infection was low (the Washington State SARS-CoV-2 test positivity rate was 3.7% to 10.2% during the study period). (12) Of the eight positive cases with either clinical follow-up or subsequent testing, five were deemed to be bona fide true positives (62.5%) and three were false positives (37.5%). This finding underscores that even tests with superlative specificity can yield a substantial percentage of false positive results under circumstances of low pretest probability. (13)

False positive results for SARS-CoV-2 have real-world implications ranging from the psychological, at the person level, to financial and societal, at the global level, stemming from workplace interruptions, travel restrictions, and misspent financial and human resources. (14) From our data, repeat testing of the same specimen on an orthogonal platform (if available) is valuable in situations with low pre-test probability. All of the false positive tests did not repeat as positive on an orthogonal platform while all true positives repeated as positive by another assay. Repeat collection and testing of another specimen, ideally within a short (several day) timeframe, can also serve to identify false positive results.

There are several limitations to the work presented here. This study was performed at a single site and our measured false positivity rate is likely dependent on our own testing processes. This analysis only covers high-throughput automated platforms. Other PCR methods that use a 96 well plate format which involve sample transfers and manipulation likely will experience a higher false positivity rate. The individuals tested here were not uniformly sampled in time after a positive result. Also, the specimen type, nasal swab, may have influenced the positive rate. Nasopharyngeal swabs are considered the gold standard, with a higher rate of positivity compared to other, less invasive sample types. (15) Specimens which resulted as negative after a prior positive were not repeated on an orthogonal platform, thus raising the specter of false negatives. It is unclear how generalizable these numbers are to other laboratories or platforms. More work is required to understand the inherent false positivity rate associated with the automated platforms in use within our laboratory.

## Data Availability

data availability statement in manuscript

## Acknowledgements

The authors acknowledge Joshua Lieberman for his helpful insights as well as the staff and patients of UW Virology and the Department of Laboratory Medicine and Pathology at the University of Washington Medical Center. ALG reports contract funding from Abbott and Gilead for testing, research funding from Merck, all outside of the submitted work.

## References

1. Burd EM. 2010. Validation of laboratory-developed molecular assays for infectious diseases. Clin Microbiol Rev 23:550–76.

2. Corman VM, Landt O, Kaiser M, Molenkamp R, Meijer A, Chu DK, Bleicker T, Brünink S, Schneider J, Schmidt ML, Mulders DG, Haagmans BL, van der Veer B, van den Brink S, Wijsman L, Goderski G, Romette JL, Ellis J, Zambon M, Peiris M, Goossens H, Reusken C, Koopmans MP, Drosten C. 2020. Detection of 2019 novel coronavirus (2019-nCoV) by real-time RT-PCR. Euro Surveill 25.

3. Ridgway JP, Shah NS, Robicsek AA. 2020. Prolonged shedding of severe acute respiratory coronavirus virus 2 (SARS-CoV-2) RNA among patients with coronavirus disease 2019 (COVID-19). Infect Control Hosp Epidemiol 41:1235–1236.

4. Plebani M. 2012. Quality indicators to detect pre-analytical errors in laboratory testing. Clin Biochem Rev 33:85–8.

5. Lin L, Carlquist J, Sinclair W, Hall T, Lopansri BK, Bennett ST. 2020. Experience with False Positive Test Results on the TaqPath Real-Time Reverse Transcription-PCR COVID-19 Testing Platform. Arch Pathol Lab Med doi:10.5858/arpa.2020-0612-LE.

6. Mögling R, Meijer A, Berginc N, Bruisten S, Charrel R, Coutard B, Eckerle I, Enouf V, Hungnes O, Korukluoglu G, Kossyvakis T, Mentis A, Molenkamp R, Muradrasoli S, Papa A, Pigny F, Thirion L, van der Werf S, Reusken C. 2020. Delayed Laboratory Response to COVID-19 Caused by Molecular Diagnostic Contamination. Emerg Infect Dis 26:1944–1946.

7. Wernike K, Keller M, Conraths FJ, Mettenleiter TC, Groschup MH, Beer M. 2020. Pitfalls in SARS-CoV-2 PCR diagnostics. Transbound Emerg Dis doi:10.1111/tbed.13684.

8. Council OotG. 2020. Summary of the Findings of the Immediate Office of the General Counsel’s Investigation Regarding CDC’s Production of COVID-19 Test Kits. Services DoHH, Washington, D.C.

9. Lieberman JA, Pepper G, Naccache SN, Huang ML, Jerome KR, Greninger AL. 2020. Comparison of Commercially Available and Laboratory-Developed Assays for. J Clin Microbiol 58.

10. Price TK, Bowland BC, Chandrasekaran S, Garner OB, Yang S. 2021. Performance Characteristics of Severe Acute Respiratory Syndrome Coronavirus 2 RT-PCR Tests in a Single Health System: Analysis of >10,000 Results from Three Different Assays. J Mol Diagn 23:159–163.

11. Gorzalski AJ, Tian H, Laverdure C, Morzunov S, Verma SC, VanHooser S, Pandori MW. 2020. High-Throughput Transcription-mediated amplification on the Hologic Panther is a highly sensitive method of detection for SARS-CoV-2. J Clin Virol 129:104501.

12. Health WSDo. 3/11/2021 2020. COVID-19 Data Dashboard, on Washington State Department of Health. https://www.doh.wa.gov/Emergencies/COVID19/DataDashboard#dashboard. Accessed 3/12/2021.

13. Lieberman JA, Lieberman SM, Bourassa LA. 2020. What tests to use, when, why—and why not? Pitfalls of mass testing for COVID-19, on USC-Brookings Schaeffer on Health Policy. https://www.brookings.edu/blog/usc-brookings-schaeffer-on-health-policy/2020/10/27/sars-cov-2-testing-what-tests-to-use-when-why-and-why-not/. Accessed 3/2/2021.

14. Surkova E, Nikolayevskyy V, Drobniewski F. 2020. False-positive COVID-19 results: hidden problems and costs. Lancet Respir Med 8:1167–1168.

15. Lee RA, Herigon JC, Benedetti A, Pollock NR, Denkinger CM. 2021. Performance of Saliva, Oropharyngeal Swabs, and Nasal Swabs for SARS-CoV-2 Molecular Detection: A Systematic Review and Meta-analysis. J Clin Microbiol doi:10.1128/JCM.02881-20.

